# An exploratory survey about using ChatGPT in education, healthcare, and research

**DOI:** 10.1101/2023.03.31.23287979

**Authors:** Mohammad Hosseini, Catherine A. Gao, David Liebovitz, Alexandre Carvalho, Faraz S. Ahmad, Yuan Luo, Ngan MacDonald, Kristi Holmes, Abel Kho

**Author notes:** Corresponding author: Mohammad Hosseini, 680 N Lake Shore Dr, Suite 1400, Chicago, IL 60611. co-first authors (Mohammad Hosseini, Catherine A. Gao).

## Abstract

**Objective:** ChatGPT is the first large language model (LLM) to reach a large, mainstream audience. Its rapid adoption and exploration by the population at large has sparked a wide range of discussions regarding its acceptable and optimal integration in different areas. In a hybrid (virtual and in-person) panel discussion event, we examined various perspectives regarding the use of ChatGPT in education, research, and healthcare.

**Materials and Methods:** We surveyed in-person and online attendees using an audience interaction platform (Slido). We quantitatively analyzed received responses on questions about the use of ChatGPT in various contexts. We compared pairwise categorical groups with Fisher’s Exact. Furthermore, we used qualitative methods to analyze and code discussions.

**Results:** We received 420 responses from an estimated 844 participants (response rate 49.7%). Only 40% of the audience had tried ChatGPT. More trainees had tried ChatGPT compared with faculty. Those who had used ChatGPT were more interested in using it in a wider range of contexts going forwards. Of the three discussed contexts, the greatest uncertainty was shown about using ChatGPT in education. Pros and cons were raised during discussion for the use of this technology in education, research, and healthcare.

**Discussion:** There was a range of perspectives around the uses of ChatGPT in education, research, and healthcare, with still much uncertainty around its acceptability and optimal uses. There were different perspectives from respondents of different roles (trainee vs faculty vs staff). More discussion is needed to explore perceptions around the use of LLMs such as ChatGPT in vital sectors such as education, healthcare and research. Given involved risks and unforeseen challenges, taking a thoughtful and measured approach in adoption would reduce the likelihood of harm.

## Introduction

The introduction of OpenAI’s ChatGPT has delivered large language model (LLM) systems to a mainstream audience. Other technologies such as Elicit, SciNote, Writefull, and Galactica, have existed, but exponential growth of ChatGPT’s audience has sparked vigorous discussions in academic circles. LLMs have demonstrated remarkable ability (and sometimes inability) in generating text in response to prompts. Some LLMs like Elicit and Med-PaLM can scan available literature and suggest specific questions or insights about a particular topic/question by leveraging available knowledge. The new GPT4 can also learn from images, thereby multiplying possible use cases of LLM, especially in education, healthcare and research settings where visual representations are fundamental to create or enhance understanding. To explore the implications of using LLMs in research, education and healthcare, Northwestern University’s Institute for Augmented Intelligence in Medicine (I.AIM) and Institute for Public Health & Medicine (IPHAM) organized a hybrid (virtual and in-person) event on Feb 16^th^ 2023 entitled “Let’s ChatGPT”. This event consisted of lively discussions and an exploratory survey of participants. In this article, we present survey results and provide a qualitative analysis of raised issues.

### Using ChatGPT and other LLMs in Education

Responses to the use of ChatGPT in education are varied. For instance, some New York schools banned students from using ChatGPT[1], while others adopted policies in their syllabus that encourage students to engage with these models as long as they disclose it[2]. Some educators fed ChatGPT questions from a freely available United States Medical Licensing Examination (USMLE) and reported a near or at passing range performance[3]. As the technology improves, the debate is still open about ethical and educational uses, with many issues remaining unresolved and concerns being explored. Among such concerns, the issue of “disguising biases” is noteworthy. It is believed that by weaving information from various sources that could be biased to generate a response, ChatGPT creates a “tapestry of biases”, thereby making it more difficult to trace the biases embedded in used sources[4].

### Using ChatGPT and other LLMs in Healthcare

There has long been excitement around the use of Artificial Intelligence (AI) in healthcare applications[5]. Language-specific applications of interest include improving efficiency of clinical documentation, decreasing administrative task burdens, creating clearer understanding for patients of complicated test result reports, and responding to in-basket Electronic Medical Record (EMR) messages. For example, Doximity released a beta version of DocsGPT, a tool that integrates ChatGPT to assist with writing clinical work such as writing insurance denial appeals.[6] There has also been exploration of using ChatGPT to answer medical questions[7], write clinical case vignettes[8], and simplify radiology reports to enhance patient-provider communication[9]. A major caveat lies in the models’ tendency to ‘hallucinate’ or ‘confabulate’ factual information, and thus, the importance of proof-reading and a domain expert reviewing and editing the output for accuracy cannot be overemphasized.

### Using ChatGPT and other LLMs in Research

Even before the introduction of OpenAI’s ChatGPT, using computer generated text in academic publications had an estimated prevalence of “4.29 papers for every one million papers” as per 2021[10]. There were concerns about the negative impact of using LLMs on the integrity of academic publications[11]. One way the community was able to detect these papers was through spotting so-called tortured phrases,(i.e., the AI-generated version of an established phrase used in specific disciplines for certain concepts and phenomena).

ChatGPT, on the other hand, generates fluent and convincing abstracts that are difficult for human reviewers or traditional plagiarism detectors to identify[12]. As ChatGPT and other recently developed applications based on LLMs mainstream the use of AI-generated content, detection will likely become much more difficult. This is partly because, (1) with an increase in the number of users, LLMs learn quicker and produce better human-like content, (2) more recent LLMs benefit from better algorithms and, (3) researchers are more aware of LLMs’ shortcomings e.g., use of tortured phrases and mix generated content with their own writing to disguise their use of LLMs. Detection applications such as the OpenAI Classifier, which uses four (ambiguous) categories to label inputted text (Very unlikely, Unlikely, Unclear if it is/Possibly or Likely AI-generated) seem unreliable and for the foreseeable future will likely remain so. Given challenges of detecting AI-generated text, it makes sense to err on the side of transparency and encourage disclosure. Various journal editors and professional societies have developed disclosure guidelines, stressing that LLMs cannot be authors[13,14], and suggesting that disclosure should happen as part of the methods section, describing who used the system, when, and using which prompts plus adding it among cited references[15]. Besides writing scholarly manuscripts, LLMs can also be used in scholarly reviews to support editorial practices,e.g., supporting the search for suitable reviewers, the initial screening of manuscripts, and the write-up of final decision letters from individual review reports, but various risks such as inaccuracies and biases require researchers to engage with LLMs cautiously[16].

## Methods

The research protocol and the first draft of survey questions were developed (M.H. and C.A.G) based on available and ongoing work about LLMs and ChatGPT, with suggestions from other panel members (K.H. and N.K.N.M) and a team member (E.W.). ChatGPT was used to brainstorm survey questions.^1^ The Northwestern IRB granted exemption (STU00218786). We received permission from the Vice Dean of Education to gather responses from medical trainees attending the session. Participants were informed about the survey details, such as anonymized data collection and voluntary participation, and were offered a chance to view the information sheet and consent form before the start of the survey. We collected anonymized data using a paid version of Slido (Bratislava, Slovakia; https://www.sli.do/). The full survey is available in the Supplemental Document.

The quantitative survey data were analyzed and visualized (C.A.G) in python v 3.8 with scipy v1.7.3, matplotlib v3.5.1, seaborn v0.11.2, tableone v0.7.10 [17], and plot_likert v0.4.0[18]. ChatGPT was used for minor code troubleshooting. For the small subset of 18 respondents who selected multiple roles, we took their most senior role and most clinical role for analysis. Binarized responses included any answer with ‘yes’, with the other category being ‘No + unsure’. Categories were compared pairwise using Fisher’s Exact tests.

The discussion was analyzed after transcribing the session (M.H.). For this purpose, we used the three topic areas highlighted in the event description (education, healthcare and research) to qualitatively code the transcripts using an inductive approach[19]. Using these codes we analyzed the transcript. Subsequently, we identified three subcodes within each code (possible positive impacts, possible negative impacts and remaining questions), bringing the total number of codes to nine. Using these nine codes, we analyzed the transcript for a second time and generated a report. Upon the completion of the first draft of the report, feedback was sought from all members of the panel and the text was revised accordingly.

## Results

### Survey results

We had 1,174 people register for the event. The peak number of webinar participants during the event was 718, and 126 people indicated they would attend in-person. We received survey responses from 420 people; a conservative estimated response rate is 49.7%. The smallest group were medical trainees (medical students, residents, and fellows) at 14 respondents (3.3% of all respondents), and second smallest by clinical faculty with 45 (10.7%) respondents (Figure 1). There were more research trainees (graduate students and postdoctoral researchers) with 53 (12.6%) respondents and research faculty with 65 (15.5% respondents). Administrative staff made up 70 (16.7%) of respondents. The largest group of respondents identified as ‘Other’, with 173 respondents (41.2% of all respondents). Full respondent breakdown and answers by respondent role are available in Table 1 of the Supplemental Document.

**Figure 1.**
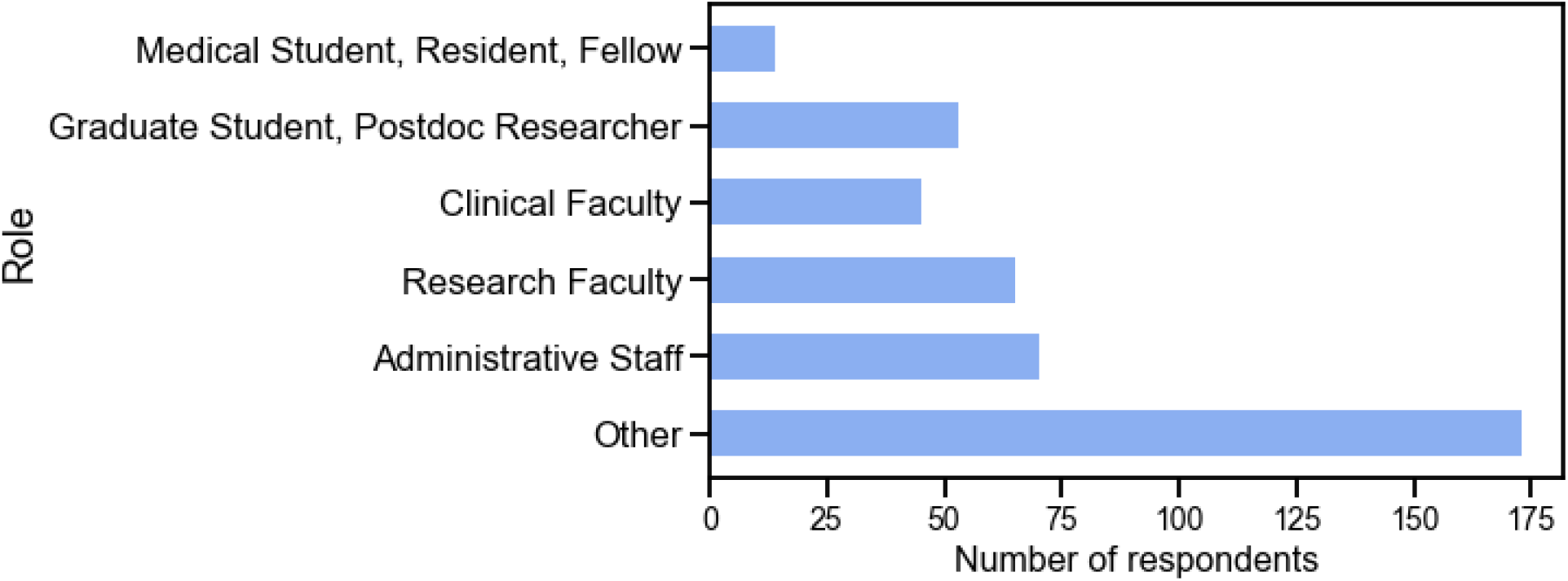
Histogram of respondents, broken down by roles.

Overall, only 40% of the audience has tried ChatGPT. Medical and research trainees were more likely to have used ChatGPT compared with faculty and staff (Figure 2). Significantly more medical trainees (medical student, residents, fellows) had tried ChatGPT (64.2%) compared with clinical faculty (31.1%), p=0.03. A more similar number of graduate students and postdoctoral researchers had tried ChatGPT (56.6%) compared with research faculty (49.2%), p=0.46.

**Figure 2.**
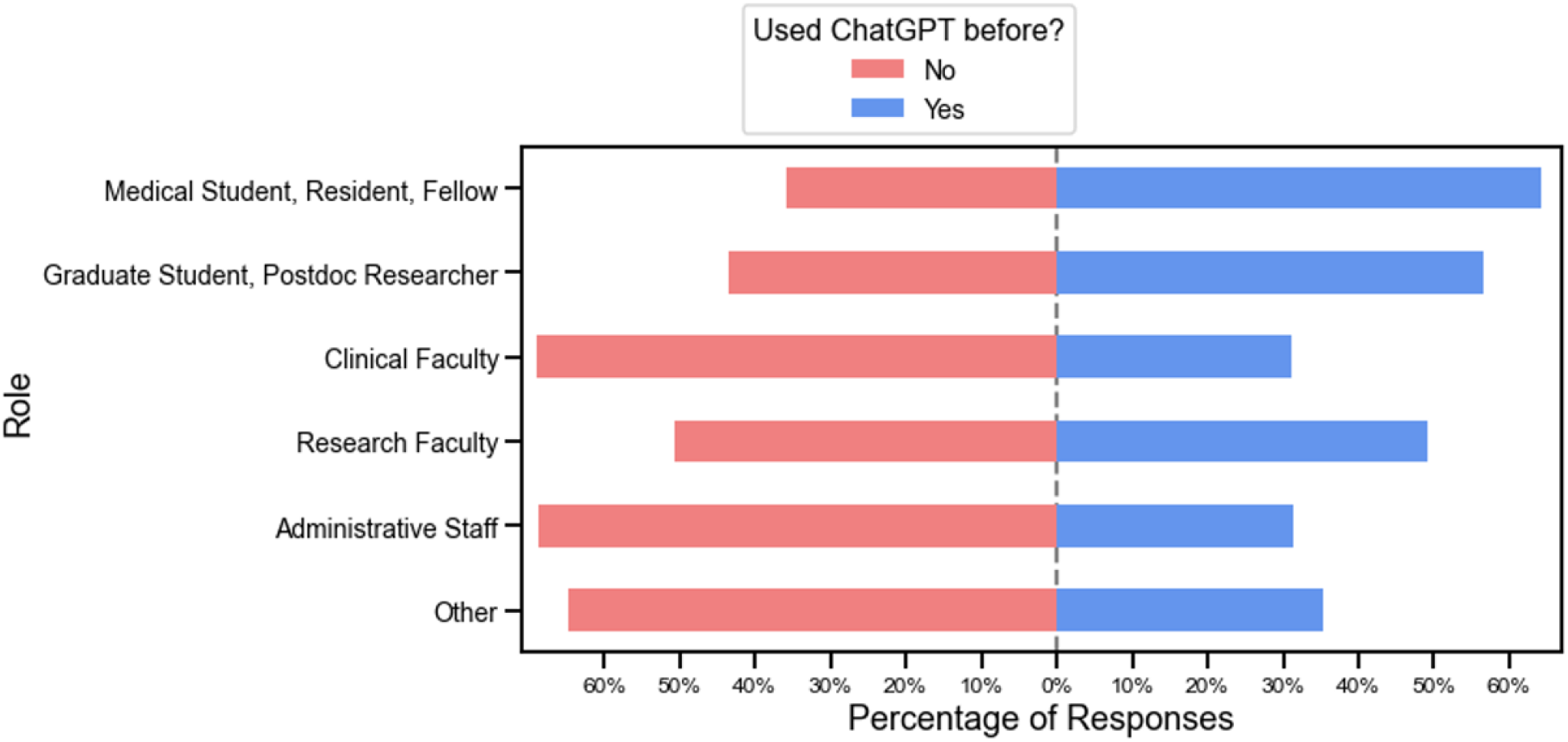
Percentage of respondents who had previously used ChatGPT, broken down by role.

Across all roles except medical trainees, the most common response regarding interest in using ChatGPT going forwards was ‘Somewhat’ (Figure 3). Those who had used ChatGPT already had higher interest in using it compared with those who did not; 39.9% had interest in using it ‘to a great extent’ compared with 15.9%, p<0.001) (Figure 4).

**Figure 3.**
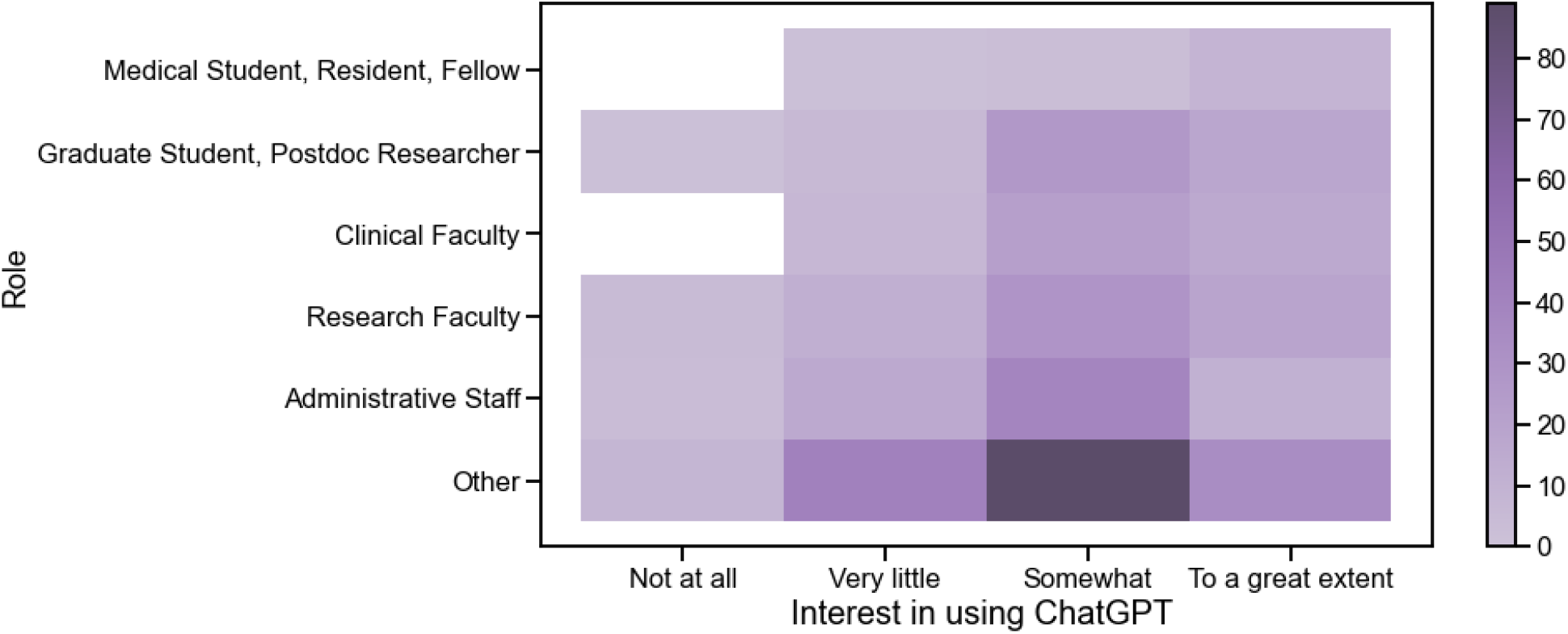
Heatmap of responses by role and interest in using ChatGPT. The most commonly selected level of interest for using ChatGPT across most roles was ‘Somewhat’.

**Figure 4.**
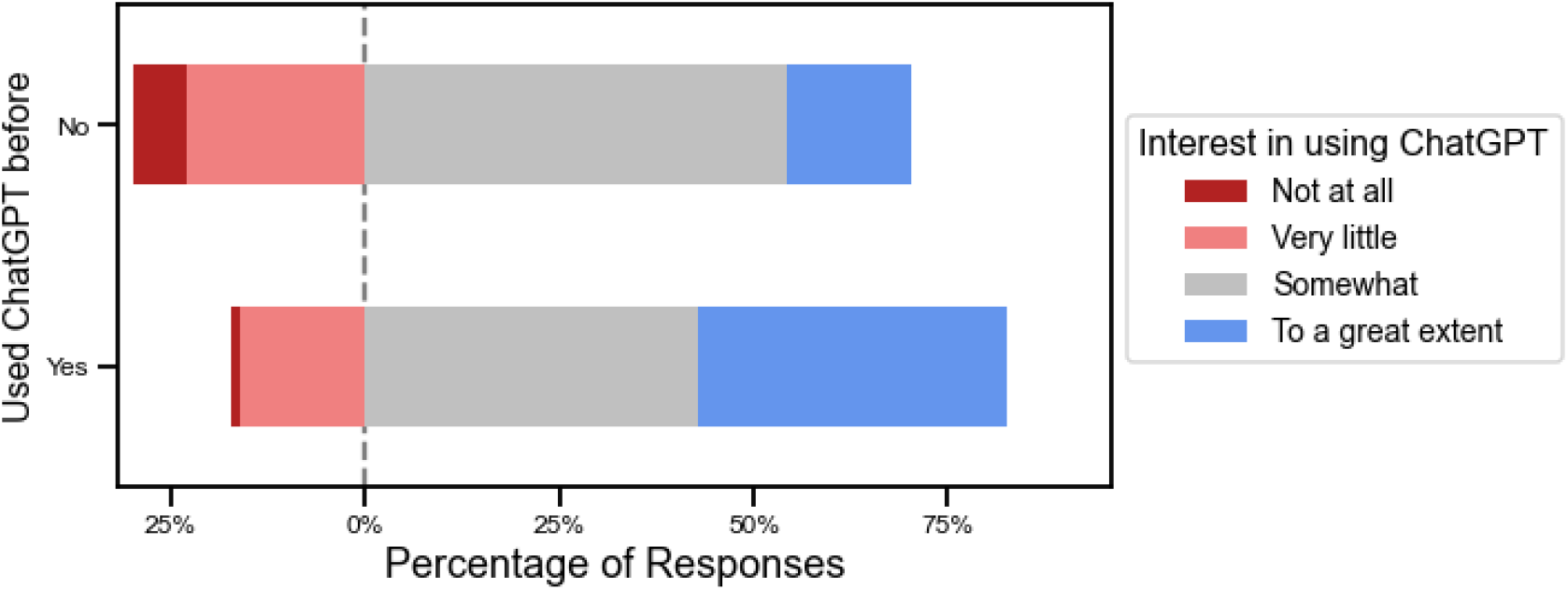
There was greater interest going forwards among those who had already tried ChatGPT compared to those who had not.

In response to questions about whether ChatGPT can be used, there was greater uncertainty around its use in Healthcare and Education, compared to using it in Research. For Research, only 75 (17.9%) of respondents selected ‘I don’t know, it is too early to make a statement’, compared with 226 (53.8%) when asked about using it in Education (p<0.001), and 177 (42.2%), when asked about using it in Healthcare (p<0.001), (Table 1, Supplemental Document). Medical and research trainees were more interested in using it for education purposes compared with clinical and research faculty, though this was not statistically significant. Of note, when responding to the question about using ChatGPT in Healthcare, a significant portion (42% of respondents) of respondents approved of using it for administrative purposes (for example, writing letters to insurance companies) and there was a smaller population of respondents who thought it could be used for any purpose (12.2%) (Table 1, Supplemental Document). Medical trainees felt it was more acceptable to use this technology for healthcare purposes (including administrative purposes), compared with clinical faculty 92.9% ‘yes’ vs 48.9% ‘yes’, p=0.004 (Figure 5).

**Figure 5.**
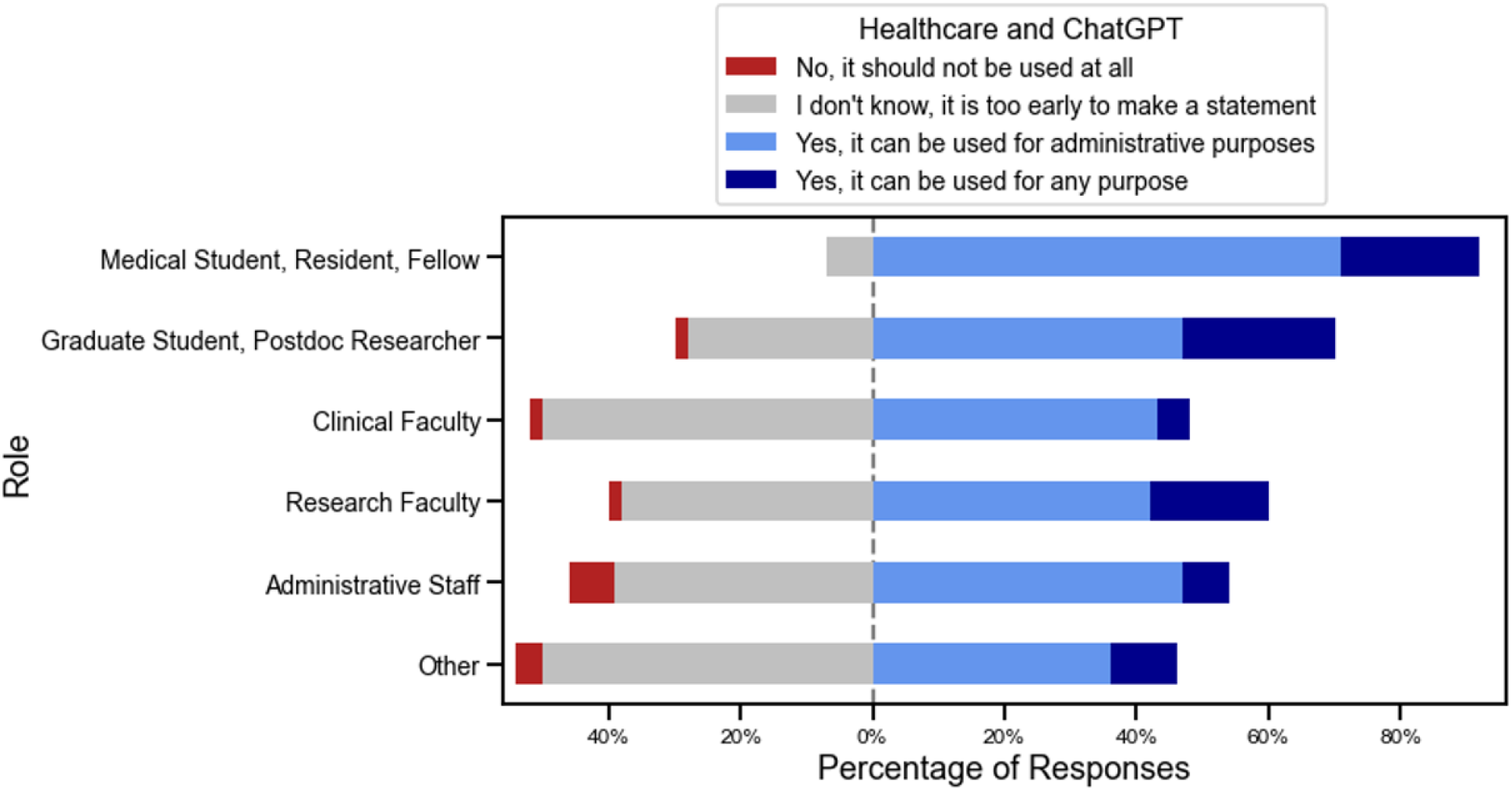
Breakdown of proportions of answers when asking about ChatGPT use in Healthcare, as split by respondent’s role.

Those who had already used ChatGPT were more likely to deem it acceptable for research purposes (89.3% ‘yes’) versus those who had not used it before (75% ‘yes’), 14.3% higher, p<0.001 (Figure 6). Similarly, those with prior experience thought it was acceptable to use in healthcare 62.5% vs 48.8%, 13.7% higher, p=0.008. They also thought it was more acceptable to use in education, 63.9% vs 30.2%, 33.7% higher, p<0.001.

**Figure 6.**
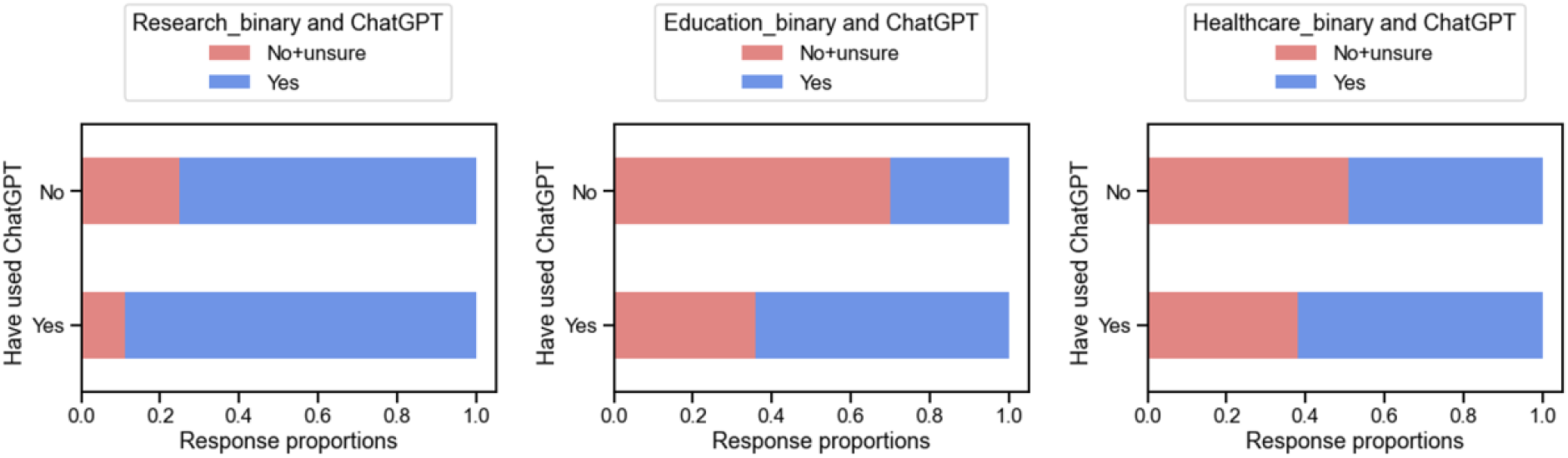
Comparison of binarized responses in use of ChatGPT for research, education, and healthcare, broken down by previous use of ChatGPT.

**Analysis of the Q&A session**

## Education

### Possible positive impacts

“Leveling the playing field” for students with different language skills was identified as an advantage of using LLMs. Since students’ scientific abilities should not be overshadowed by their insufficient language skills, ChatGPT was seen as a solution that could help fix errors in writing and accordingly, an instrument that can support students who might be challenged by writing proficiency — specifically those not writing in their native language. Another useful application was “adding the fluff” to writing (i.e., details that could potentially improve comprehension), especially for those with communication challenges. Structuring and summarizing existing text or creating the first draft of letters of application with specific requirements were also mentioned among possible areas where ChatGPT could help students. Another mentioned possibility was to use ChatGPT as a studying tool that (upon further improvements and approved accuracy) could describe specific medical concepts at a specific comprehension level (e.g., “explain tetralogy of fallot at the level of a tenth grader”).

### Possible negative impacts

Given existing inaccuracies in systems such as ChatGPT, a panel member warned medical students against using them to explain medical concepts and were encouraged to have everything “double and triple checked”. To the extent that ChatGPT could be used to find fast solutions, and as a substitute for hard work and understanding the material (e.g., only to get through the assignments or take shortcuts), it was believed to be harmful for education. Clinical-reasoning skills were believed to be at risk if ChatGPT-like systems are used more widely. For instance, it was believed that writing clinical notes helps students “internalize the clinical reasoning that goes into decision making”, and so until such knowledge is cemented, using these systems would be harmful for junior medical students. One member of the audience warned that since effective and responsible use of ChatGPT requires adjusted curricula and assessment methods, employing them before these changes are enacted would be harmful. A panel member highlighted the lack of empirical evidence in relation to the usefulness and effectiveness of these systems when teaching different cohorts of students with various abilities and interests. As such, early adoption of these systems in all educational contexts was believed to have unforeseen consequences.

### Remaining questions

Challenges of ensuring academic integrity and students’ willingness to disclose the use of ChatGPT were raised by some attendees. However, as a clinical faculty member suggested, these are neither new challenges nor unique problems associated with ChatGPT because even in the absence of such tools, one could hire somebody to write essays. Plagiarism detection applications and stricter regulations have not deterred outsourcing essay writing. Therefore, it remains an open question as to how ChatGPT changes this milieu.

A panelist suggested that similar to when ChatGPT is used to write code (e.g., in Python) and the natural tendency to test generated code to see if it actually works (e.g., as part of the larger code), students should employ methods to test and verify the accuracy and veracity of generated text. However, since systems like ChatGPT are constantly evolving, developing suggestions and guidelines for verification is challenging.

Information literacy was another issue raised by a panelist. New technologies such as ChatGPT extend and complicate existing discussions in terms of how information is accessed, processed, evaluated and ultimately consumed by users. From a university library perspective, training and supporting various community members to responsibly incorporate new technology in decision making and problem solving requires mobilizing existing and new resources.

## Healthcare

### Possible positive impacts

Improving communication between clinicians and patients was among possible gains. For example, it was highlighted that “doctors might not be in their best self” during an extremely busy week when they are responding to patient’s EMR messages, and so ChatGPT could ensure that all niceties are there, include additional content based on patients’ history and maintain emotional consistency in communication. Upon further development, these systems could help centralize and organize patient records by flagging areas of concern to improve diagnosis and effective decision making. Currently, our medical records lack sufficient usability and when assessing patients, one is concerned that some vital information might be “buried in a chart” that is not readily accessible, with LLMs acting as “assistants” or “co-pilots”, able to find these hidden and sometimes critical pieces of information for the provider saving time and improving care delivery

Efficiency of documentation was highlighted as an important gain for clinicians, patients and the healthcare system. For example, increased efficiency in note-taking through prepopulation of forms, voice recording and morphing that into clinician notes, and synthesizing existing patient notes to save clinicians’ time were noted as possibilities. This increased efficiency was believed to benefit patients through improved care and increased patient-clinicians interaction time, which could improve shared decision-making conversations. One panelist highlighted that patient notes are logged in the EHR system mostly late at night or during off hours, stressing the burden of note taking on clinicians as a driver of burnouts.

### Possible negative impacts

Given recent evidence about ChatGPT’s inaccuracies and so-called hallucinated content[20] as well as lack of transparency about used sources in training it, using these systems in triage and preparing for new patients or for clinical diagnosis was deemed risky. One panelist highlighted previous failures of AI models in clinical settings [21,22] as a lesson for the community to adopt these technologies with caution and only after regulatory approvals. Furthermore, the COVID-19 pandemic and clinicians’ experience of having to fight “malicious misinformation” was used as an example to highlight risks associated with irresponsible use. Malevolently using wrong or inaccurate data to train an LLM was described as “poisoning the dataset” to produce a predictive model that generates erroneous information.

Although the speculated positive impact on efficiency was mostly seen positively, some shared reservations about it, highlighting that the freed-up time could be seen as an opportunity to ask clinicians to visit more patients instead of spending more time with them. The explanation was that the healthcare system could redirect an opportunity like this to generate additional revenue. Furthermore, using technology to consolidate existing notes or pre-populate forms was believed to increase the likelihood that falsehood could be copy-pasted and result in carrying forward errors. The concern being that since these systems have the propensity to pass on information *as well as misinformation*, wrong diagnoses could be carried forward without being questioned. Unless the veracity of carried historical information is questioned, clinicians might be trained out of the habit of critical thinking and assume all information as reliable.

### Remaining questions

In discussing incorporation of ChatGPT in healthcare, specific techno-ethical challenges were highlighted. It was stressed that while excitement about technology is positive, specific aspects need profound deliberation and intentional design. These include defining and enforcing different access levels (e.g., to clinical notes), regulating data reuse, protecting patients’ privacy, accountability of user groups, and credit attribution for data contributions. Furthermore, securing the required financial investment to incorporate LLMs into existing IT infrastructure and workflows was believed to be challenging.

Upon debating as to whether ChatGPT is a friend or foe, one panelist mentioned challenges such as distribution disparities, and said “unfortunately, the track record of our use of technologies is not strong. New technologies have always worsened disparities and I have a significant concern that the computer power that is needed to generate and power these systems will be inadequately distributed.”

When discussing the risk of malevolently poisoning LLMs’ training data, one panelist highlighted that it remains unclear how LLMs’ healthcare data should be curated and how erroneous information could be identified and removed. Furthermore, who should be responsible to monitor the sanctity of training data or prioritize available information (e.g., based on the reliability of used sources)? It was noted that when using sources such as Google, users have already developed specific skills to question unique sources but because ChatGPT “assimilates” enormous amounts of information, attributions are ambiguous and so verification remains challenging.

## Research

### Possible positive impacts

Refining scholarly text or making suggestions to improve existing texts were highlighted among possible positive impacts. Support provided by a writing center were used as an analogy to describe some of these gains. One unique feature of ChatGPT was believed to be bidirectional communication, which allows (expert) users to “interrogate the system and help refine the output”, which will ultimately benefit all users in the long run.

### Possible negative impacts

Lack of transparency about the used data to train LLMs was believed to hide biases and disempower researchers in terms of “grasping the oppression that has gone into the answers”. This issue was also stressed by a member of the audience who questioned the language of used sources. One panelist speculated that the training data likely contained more sources in overrepresented languages within the scholarly corpus (e.g., English, French). Furthermore, since ChatGPT is currently made unavailable (by OpenAI) in countries such as China, Russia, Ukraine, Iran and Venezuela, it cannot be trained by or receive feedback from researchers who are based in these countries, and thus, might be biased towards the views of researchers based in specific locations.

### Remaining questions

One member of the audience believed that disclosure guidelines (e.g., researchers to disclose what part of the text is influenced by ChatGPT) are unenforceable and so, their promotion is moot. They added that the existing norms on plagiarism cover potential misconduct using ChatGPT. One panelist agreed with the unenforceability of guidelines (because researchers may alter AI-generated text to disguise their use), but highlighted that given the novelty of ChatGPT and its unique challenges, good practices in relation to this technology should be specified and promoted nonetheless.

#### Word cloud

We asked attendees to describe the most important risks and benefits of using ChatGPT with only one keyword. After correcting typos and replacing all plurals with singular words (with the help from ChatGPT), we used a free online word cloud generator (https://www.jasondavies.com/wordcloud/) to produce the following two figures (Figure 7A and 7B).

**Figure 7.**
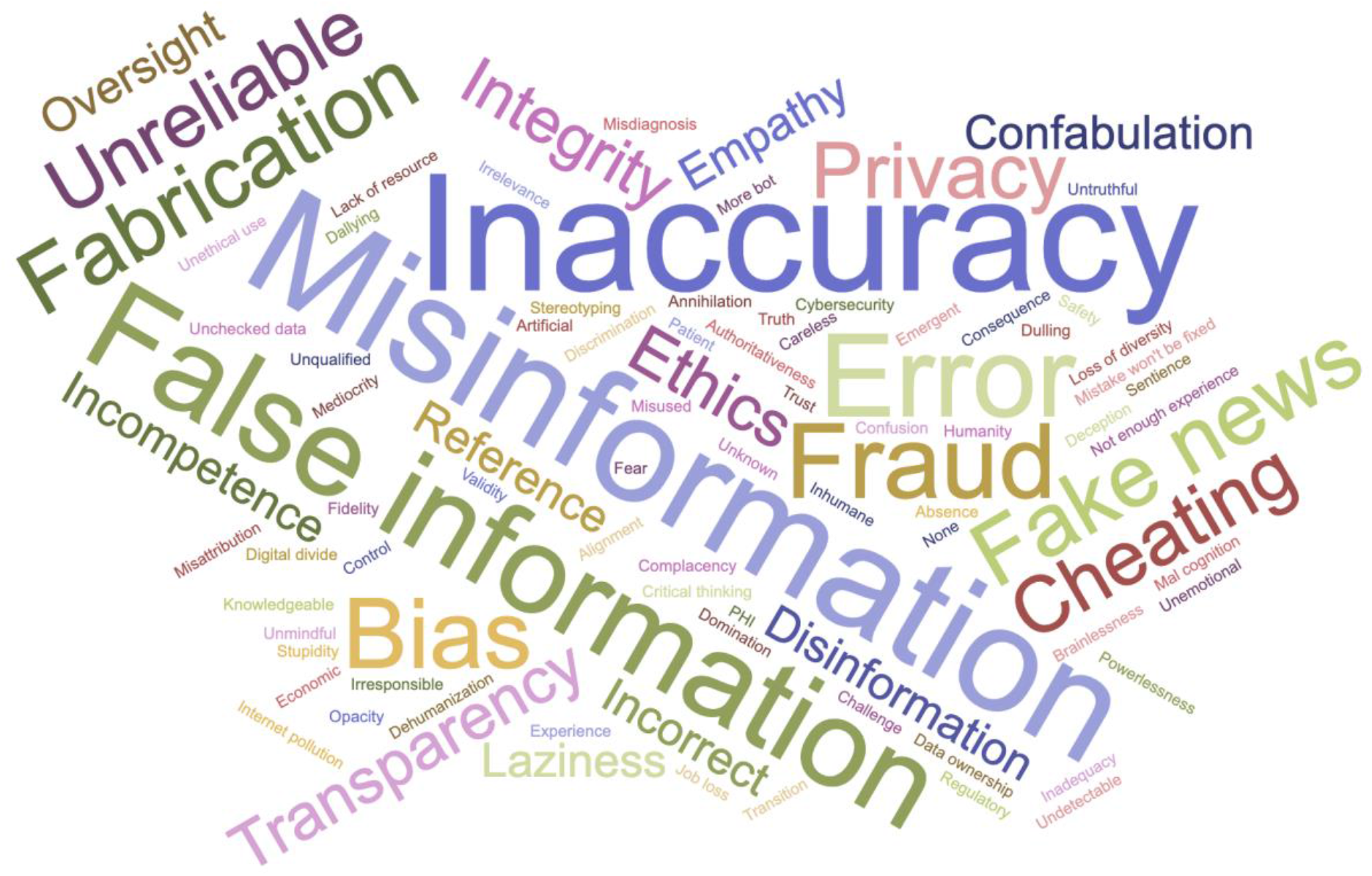
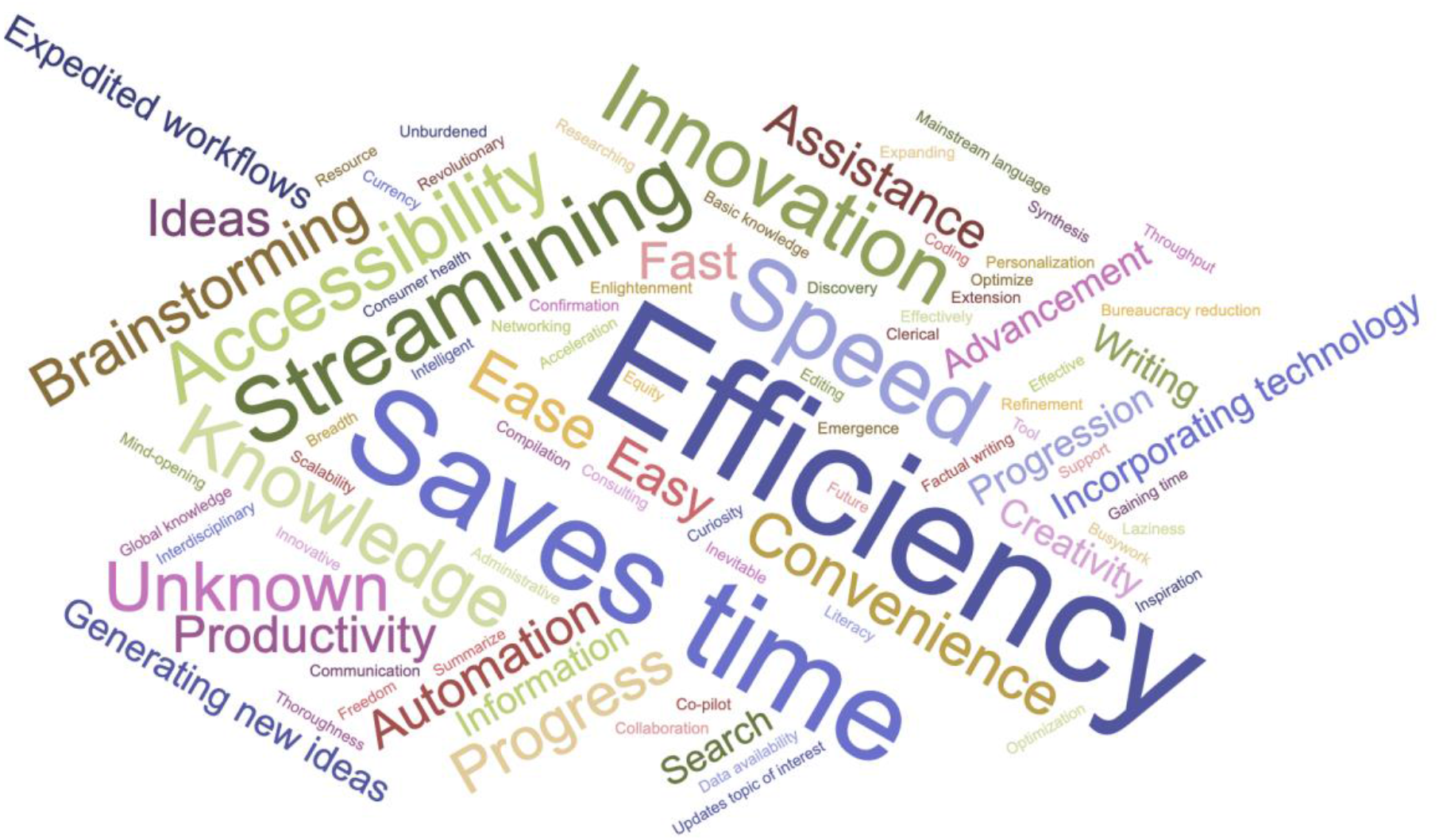
(A). With one keyword, describe the most important risk of using ChatGPT (n=225). (B) With one keyword, describe the most important benefit of using ChatGPT (n=263).

## Discussion

We hosted a large forum to explore perspectives on the interest and use of ChatGPT across education, research, and healthcare purposes. Overall, there was still a lot of uncertainty around the acceptability of its use, with a large portion of respondents saying it was too early to make a statement and that they remained somewhat interested in using ChatGPT. Trainees were more interested in using ChatGPT than faculty, having more positive views, interest, and acceptability beliefs in using the technology. More trainees than faculty had already tried ChatGPT. This points to a potential generational divide between early adopters (trainees) and late adopters (faculty), with the latter in positions of power to dictate policy to trainees and the academic community at large. Therefore, it is important that shared decision-making about appropriate use incorporates the voices of all stakeholders, with emphasis on the input of trainees, which are the ones most likely to be impacted by the continued development and deployment of this nascent technology. There was greater consensus that it was acceptable to use for research and healthcare (including for administrative tasks) than there was for education purposes. Policies may be helpful in clarifying what is deemed acceptable use, so as to avoid miscommunication or ambiguity. Future studies could examine each arena in greater detail, specifically among the population of potential users.

Exploration of the technology should be encouraged. Only 40% of our respondents had already tried ChatGPT. Participants that had used the technology before had a tendency to have a more optimistic outlook about LLMs in general whereas never-users seemed to have more concerns about its widespread adoption. Thus, it is important to continue to educate and inform the population about LLMs and their responsible use through practical applications (including live demonstrations), so never-users can grasp the technology and help dispel the fear of the unknown and promote equity. Using and engaging with LLMs is essential to learning their abilities and limitations.

Respondents and audience members had a wide range of interesting points with regards to the use of ChatGPT for research, education, and healthcare, with a mixture of positive and negative responses. Ongoing discussion is essential, especially given the current “black-box” nature of ChatGPT, with users left in the blind on *how* the LLM produced its outputs to users prompts.

Unresolved questions remain about how it curates content, the corpus of data it is trained on, the weights it uses to sort out evidence, and the risks of spreading fake news, misinformation or bias. One potential solution from legislators would be to require increased transparency from OpenAI and other LLM companies.

## Limitations

Some of the limitations include inability to break down and better delineate the large “Other” category of respondents. Since respondents were likely interested in ChatGPT to register for and attend the event, and also complete the survey, our results might not be representative of the various cohorts within the academic community.

Although medical trainees had positive views towards ChatGPT and its use, they were our smallest group of respondents (3.3% of our cohort). We took a neutral tone to the technology in our recruitment material for the event, as evidenced by the respondents from other roles who had more lukewarm or uncertain feelings towards ChatGPT. Hence, we suspect there is high interest from this group inherent to their role. Future studies could focus more closely on examining this group in particular.

## Conclusion

There is still much to discuss about the optimal and ethical uses of LLMs such as ChatGPT. Responsible use should be promoted by all, and future discussion should continue to explore the boundaries of this technology. LLMs and AI in general have the potential to change the fabric of society and impact labor relations at large, deeply transforming *how we relate to one another and do work*. However, it seems to be a double-edged sword, bringing with it the promise of more efficiency, creativity and free time for all, but risking spreading bias, hate, misinformation, and furthering the digital divide between people that have access to technology and are fluent in its use versus the ones left behind. The broad interest and engagement sparked by ChatGPT strongly suggests that, while a work in progress, LLMs have a significant potential for disruption. To navigate this uncharted territory of artificial intelligence, we recommend that future explorations of its responsible use be grounded in principles of transparency, equity, reliability, and above all, *primum non nocere*.

## Data Availability

Data availability: Data are available upon reasonable request to the corresponding author.
Code availability: Code used in this analysis and visualization are available at https://github.com/cloverbunny/gptsurvey/blob/main/gptsurvey.ipynb.

https://zenodo.org/record/7789186#.ZCb0eezML0o

## Acknowledgements

The authors wish to thank and acknowledge Eva Winckler for her contributions to event organization and coordination.

## Statements

### Data availability

Data are deposited https://zenodo.org/record/7789186#.ZCb0eezML0o, and are available upon submitting reasonable requests.

### Code availability

Code used in this analysis and visualization are available at https://github.com/cloverbunny/gptsurvey/blob/main/gptsurvey.ipynb.

## Author contributions

All authors read and approved the final draft of the manuscript. Individual contributions, as per CRediT roles:

Conceptualization: CAG; MH

Data Curation: CAG; MH

Formal analysis: CAG

Funding acquisition: AK

Investigation: CAG; MH

Methodology: CAG; MH

Project administration: CAG; MH

Software: CAG

Supervision: AK; DL; KH

Validation: AK; DL; FSA; KH; NM; YL

Visualization: CAG

Writing - Original Draft: AC; CAG; MH

Writing - Review & Editing: AC; AK; CAG; DL; FSA; KH; MH; NM; YL

## Footnotes

1 D.L. used OpenAI ChatGPT on 27th of January 2023 at 6:06pm CST using the following prompt: “please create survey questions for medical students, medical residents, and medical faculty members to answer regarding ideas for use and attitudes surrounding use of ChatGPT in education and research” (OpenAI ChatGPT, 2023).

