## Supplemental Document for "An exploratory survey about using ChatGPT in education, healthcare, and research"

*Table 1. Respondents' selections, grouped by respondent role.*

| | | | **Grouped by Role** | | | | | |
|---|---|---|---|---|---|---|---|---|
| | | Overall | Medical Student, Resident, Fellow | Graduate Student, Postdoc Researcher | Clinical Faculty | Research Faculty | Administrative Staff | Other |
| **n** | | 420 | 14 | 53 | 45 | 65 | 70 | 173 |
| **Used_before, n (%)** | No | 252 (60.0) | 5 (35.7) | 23 (43.4) | 31 (68.9) | 33 (50.8) | 48 (68.6) | 112 (64.7) |
| | Yes | 168 (40.0) | 9 (64.3) | 30 (56.6) | 14 (31.1) | 32 (49.2) | 22 (31.4) | 61 (35.3) |
| **Interested, n (%)** | Not at all | 19 (4.5) | | 2 (3.8) | | 5 (7.7) | 4 (5.7) | 8 (4.6) |
| | Very little | 85 (20.2) | 2 (14.3) | 6 (11.3) | 7 (15.6) | 12 (18.5) | 16 (22.9) | 42 (24.3) |
| | Somewhat | 209 (49.8) | 3 (21.4) | 27 (50.9) | 22 (48.9) | 29 (44.6) | 39 (55.7) | 89 (51.4) |
| | To a great extent | 107 (25.5) | 9 (64.3) | 18 (34.0) | 16 (35.6) | 19 (29.2) | 11 (15.7) | 34 (19.7) |
| **Education, n (%)** | No, it should be banned | 11 (2.6) | 1 (7.1) | | 1 (2.2) | 1 (1.5) | 2 (2.9) | 6 (3.5) |
| | I don't know, it is too early to make a statement | 226 (53.8) | 4 (28.6) | 23 (43.4) | 24 (53.3) | 36 (55.4) | 39 (55.7) | 100 (57.8) |
| | Yes, it should be actively incorporated | 183 (43.6) | 9 (64.3) | 30 (56.6) | 20 (44.4) | 28 (43.1) | 29 (41.4) | 67 (38.7) |

| | | | | | | | | |
|---|---|---|---|---|---|---|---|---|
| **Research, n (%)** | No, it should not be used at all | 6 (1.4) | | | | 3 (4.6) | 1 (1.4) | 2 (1.2) |
| | I don't know, it is too early to make a statement | 75 (17.9) | 1 (7.1) | 4 (7.5) | 12 (27.3) | 6 (9.2) | 14 (20.0) | 38 (22.1) |
| | Yes, but it should only be used to help brainstorm | 68 (16.3) | | 17 (32.1) | 5 (11.4) | 8 (12.3) | 10 (14.3) | 28 (16.3) |
| | Yes, as long as its use is transparently disclosed | 259 (62.0) | 12 (85.7) | 28 (52.8) | 26 (59.1) | 46 (70.8) | 44 (62.9) | 103 (59.9) |
| | Yes, disclosure is NOT needed | 10 (2.4) | 1 (7.1) | 4 (7.5) | 1 (2.3) | 2 (3.1) | 1 (1.4) | 1 (0.6) |
| **Healthcare, n (%)** | No, it should not be used at all | 15 (3.6) | | 1 (1.9) | 1 (2.3) | 1 (1.5) | 5 (7.1) | 7 (4.0) |
| | I don't know, it is too early to make a statement | 177 (42.2) | 1 (7.1) | 15 (28.3) | 22 (50.0) | 25 (38.5) | 27 (38.6) | 87 (50.3) |
| | Yes, it can be used for administrative purposes | 176 (42.0) | 10 (71.4) | 25 (47.2) | 19 (43.2) | 27 (41.5) | 33 (47.1) | 62 (35.8) |
| | Yes, it can be used for any purpose | 51 (12.2) | 3 (21.4) | 12 (22.6) | 2 (4.5) | 12 (18.5) | 5 (7.1) | 17 (9.8) |

*Survey delivered via Slido.*

1. **What's your current role?**
    - Medical Student, Resident, Fellow
    - Graduate Student, Postdoc Researcher
    - Clinical Faculty
    - Research Faculty
    - Administrative Staff
    - Other
2. **Have you used ChatGPT?**
    - Yes
    - No
3. **How interested are you in using ChatGPT in your day to day work?**
    - To a Great Extent
    - Somewhat
    - Very Little
    - Not at All
4. **Can ChatGPT be used in education?**
    - No, it should be banned
    - Yes, it should be actively incorporated
    - I don't know, it is too early to make a statement
5. **Can ChatGPT be used for science?**
    - No, it should not be used at all
    - Yes, but it should only be used to help brainstorm
    - Yes, as long as its use is transparently disclosed
    - Yes, disclosure is NOT needed
    - I don't know, it is too early to make a statement
6. **Can ChatGPT be used in healthcare?**
    - No, it should not be used at all
    - Yes, it can only be used to help write administrative content such as emails to insurance companies or to patients
    - Yes, it can be used for any purpose
    - I don't know, it is too early to make a statement
7. **Using one keyword, describe challenges of using ChatGPT**
8. **Using one keyword, describe benefits of using ChatGPT**
